# COVID-19 Acceleration and Vaccine Status in France - Summer 2021

**DOI:** 10.1101/2021.09.18.21263773

**Authors:** Christelle Baunez, Mickael Degoulet, Stéphane Luchini, Patrick A. Pintus, Miriam Teschl

## Abstract

**Objectives:** This note provides an assessment of COVID-19 acceleration among groups with different vaccine status in France.

**Methods:** We assess viral acceleration using a novel indicator introduced in Baunez et al. (2021). The acceleration index relates the percentage change of tests that have been performed on a given day to the percentage change in the associated positive cases that same day. We compare viral acceleration among vaccinated and unvaccinated individuals in France over the period May 31st - August 29, 2021.

**Results:** Once the state of the epidemic within each groups is accounted for, it turns out that viral *acceleration* has since mid-July converged to similar levels among vaccinated and unvaccinated individuals in France, even though viral *speed* is larger for the latter group compared to the former.

**Conclusion:** Our results call for an increasing testing effort for *both* vaccinated and unvaccinated individuals, in view of the fact that viral circulation is currently accelerating at similar levels for both groups in France.

## 1 Introduction

Existing vaccines against SARS-CoV-2 currently on the market in Western societies are said to have a high efficacity in reducing the number of symptomatic cases, and also in cutting transmission (e.g. Layan et al. 2021, Olliaro et al. 2021, Prunas et al. 2021, Salo et al. 2021). But the Delta variant of SARS-CoV-2 has created new uncertainties notably with respect to transmission and vaccin breakthrough, which refers to the observation that even vaccinated individuals may get infected and also transmit the virus to other individuals (e.g. Gazit et al. 2021, Riemersma et al. 2021, Brown et al. 2021). A certain number of studies are starting to suggest that the Delta variant “[…] is believed to spread faster than other variants” (Planas et al. 2021) and the US-based Centers for Disease Control and Prevention (CDC) indicates for example that fully vaccinated people with the Delta variant can spread the virus, although it appears for a shorter period of time.^1^ It is therefore of great importance for any public health authority to be able to measure and understand correctly the viral spread, including among the vaccinated population.

In France, we observe that at the end of August 2021, about 6 times more new positive cases are found among the unvaccinated individuals, compared to the vaccinated. This indicates that new infections are much larger for the unvaccinated but those absolute levels do not correct for the fact that unvaccinated people are tested much more each day (about 3 times more, as of August 29, 2021). The usual way to account for tests is to compute the ratio of the number of new positive cases generated on a given day to the number of new tests performed on that same day, the positivity rate. At the end of August, the positivity rate for the unvaccinated about is twice as large compared to that of the vaccinated.^2^ This suggests that the virus is spreading faster among the non-vaccinated than the vaccinated population and one may conclude that vaccines have succesfully contributed in slowing down viral transmission among the vaccinated group.

The challenge with this conclusion however is that a direct comparison between the positivity rate of different groups is not as straightforward. One general difficulty is that it relies on the implicit assumption that those positivity rates do not change with sample size. If, for instance, the number of tests of vaccinated would be increased to match those of non-vaccinated, their positivity rate is likely to differ. One reason for this is the question of who is getting tested. If among vaccinated people primarily those with symptoms are getting tested, nothing can be inferred about whether there may be asymptomatic cases among vaccinated people. Current research suggests that asymptomatic vaccinated may have a lower viral charge than asymptomatic unvaccinated individuals, and would thus be contributing less to transmission, but the full verdict is not yet known (Blanquart et al. 2021, Chau et al. 2021, Riemersma et al. 2021). Another important difficulty is that the positivity rate is level-dependent. A more meaningful comparison however puts the new share of tests that return a positive result in relation to the cumulated level of tests and cases in order to see how the daily positivity rate evolves with respect to its history in the respective group. This makes the measure level-independent in a way that indicates whether the viral spread is accelerating or decelerating, which is, as we argue below, more important information for public health than any variations of viral speed over time.

In Baunez et al. [1], we propose an acceleration index that monitors, in real-time, whether the pandemic is decelerating or accelerating, that is, whether harm (here represented as the number of cases) is accelerating or decelerating (Taleb 2012).The advantage of this index is that it measures the sensitivity of cases with respect to a change in tests. The intuition is that if we increase the amount of tests by a certain percentage and we find a greater positive percentage change of cases, then the pandemic is accelerating. If we find less than that percent change of cases, the pandemic is decelerating. The index is thus an elasticity, which measures the responsiveness or sensitivity of one variable (here cases) in response to another variable (here tests). It gives a number that takes account of the level of tests and cases and is, consequently, more informative and precise than the positivity rate about viral propagation. As long as acceleration is greater than one, the pandemic is *not* under control, whether the daily positivity rate is small or not. For a succesful vaccination campaign, we want to see low speed *and* deceleration of viral spread. If the viral transmissions are low, but with a tendency to accelerate, vaccine campaigns alone will not be able to curb the pandemic. This translates directly into an important policy goal: to curb the pandemic, it is important to test ever more, even alongside a vaccine campaign, and to find ever fewer cases. That is, the policy goal should be deceleration, and not only lower speed.

In this paper, we calculate the acceleration index for the vaccinated and unvaccinated population since the start of the vaccination campaign in France until the end of current available data, which is August, 29 2021. What we find is that despite vaccination, the pandemic is still accelerating in both population groups, although we see a decrease in acceleration in both groups since about the beginning of August. Moreover, despite lower absolute numbers among the vaccinated, viral spread accelerates similarly among the vaccinated and non-vaccinated populations. This finding suggests that vaccines are not fully protective against infection and cannot, by themselves alone, curb the pandemic especially if even vaccinated people continue to transmit the virus. Testing remains an important instrument to observe viral spread together with additional health policies such as, for example, contact tracing, quarantine, mask-wearing and social distancing, if necessary.

## 2 Methods

In Baunez et al. [1], we have proposed a real-time indicator, the acceleration index, that measures whether the pandemic is accelerating or decelerating in a given population. The acceleration index is defined as the elasticity of cumulated positive cases to cumulated tests. It is thus a level-free measure of responsiveness of cases to tests in relative terms. Suppose that data is available about the number of tested and positive persons, up to end date *T*. Denote {*p*_1_, …, *p*_*T*_} the historical times series of the new (per period) number of positive persons from date *t* = 1 to end date *t* = *T*. Similarly, {*d*_1_, …, *d*_*T*_} is the historical times series of new (per period) diagnosed/tested persons. Denote 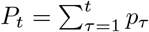 and 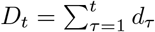 the cumulative numbers of positive and diagnosed persons up to date *t*. Importantly, variables *P*_*t*_ and *D*_*t*_ therefore define the state of the epidemic at date *t*, in terms of the current stocks of positives and tests, cumulated since date 1. In fact, those states reflect the history of each variable and it might differ across the groups of individuals that we consider below, due to vaccine status.

The acceleration index, denoted *ε*_*T*_ at date *T*, thus gives the percentage change of cases divided by the percentage change of testing as follows:

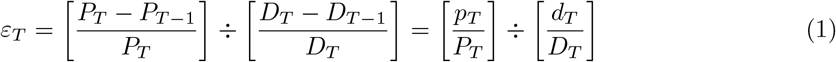

where the latter equality follows from the very definition of cumulated variables, the variations of which are just daily flows. When *ε*_*T*_ *>* 1 we say that the epidemic is *accelerating* (it is on the loose), since a given growth rate of cumulated tests produces a larger growth rate of cumulated positive cases, while it is *decelerating* (the pandemic is under control) when *ε*_*T*_ < 1. As a consequence, our indicator can tightly be linked to an arguably desirable objective of public health policy, which is to get proportionally less infected people when tests are increasing.

Rearranging the terms of the latter equality, we see that the acceleration index relates to the daily and average positivity rates, in the following way:

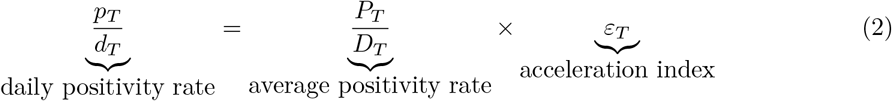

where the average positivity rate is here defined as the ratio of stocks at end date *T*. In sum, the acceleration index is the elasticity of cumulated positive cases to cumulated tests (that is, the ratio of their growth rates), which may also be decomposed as the ratio between daily and average positivity rates. It then follows that if the daily positivity rate is always constant, then *ε*_*T*_ = 1 because current and average speeds are equal. If *ε*_*T*_ *>* 1, daily positivity rate exceeds average rate, indicating acceleration, while deceleration prevails when *ε*_*T*_ < 1. We compute the acceleration index for France over the period May 31st - August 29, 2021, for individuals who are grouped according to their vaccine status. We use data about vaccine status in France that exploit two French databases SI-DEP and VAC-SI. SI-DEP contains, for each day, the total number of PCR tests realised as well as the number of positive tests. VAC-SI contains the vaccine status of the tested patients. Vaccine status is divided into four categories: (1) not vaccinated, (2) one vaccine jab for less than 7 days for Pfizer, Moderna and Astra Zeneca vaccines or 14 days for Janssen, (3) one vaccine jab for more than 7 days or two jabs for less than 7 days (Pfizer, Moderna, Astra Zeneca) and (4) full vaccination (2 jabs for more than 7 days for Pfizer, Moderna and Astra Zeneca and jab for more than 14 days for Janssen. The matching has been done by the DREES (Direction de la recherche, des études, de l’évaluation et des statistiques) from the Ministry of Health (DREES, 2021) and data are open source^3^.

We also estimate a Pearson correlation coefficient on a rolling window of 7 days between the acceleration index for the fully vaccinated and that of the recent incomplete vaccination, efficient incomplete vaccination, not vaccinated. This provides a simple measure of moment-by-moment local synchrony of viral propagation across groups with different vaccine status.

## 3 Results and Discussion

Our main results are depicted in Figure 1. Panel (*a*) depicts the amount of daily tests that are performed for the 4 groups. It reveals that the amount of testing is both time-varying and dependent on vaccine status, with unvaccinated individuals getting tested about 3 to 5 more times over the considered period. Panel (*b*) in Figure 1 depicts the daily positivity rates, defined as *p*_*T*_*/d*_*T*_ at each end date *T*, for the four groups. Looking at daily positivity rates alone, one concludes that viral speed is greater among unvaccinated people, particularly when compared with fully vaccinated individuals. On August 29, the daily positivity rate for unvaccinated people is about twice as large compared to that of the fully vaccinated. However comparing levels of the daily positivity rate across groups does not give any indication about whether the pandemic worsens faster for the unvaccinated or improves quicker for the vaccinated. For this to know, one needs to look at the acceleration of viral spread for both groups. This means to look at the change of speed or viral spread, which can be understood by putting the daily positivity rate in relation to the average positivity rate. Doing so, one obtains a level-independent measure of the dynamic of the pandemic because it puts the change in viral spread expressed through the daily positivity rate in relation with the “past” viral spread or average positivity rate, that is the cumulated number of cases over cumulated number of tests. Intuitively this means that if today we find more cases per tests than we did on average on all previous days, then the pandemic is accelerating, if we find less cases per tests today than we did on average in the past, the pandemic decelerates. This measure thus is a more informative way to compare between groups because it tells us that given current testing strategies, if of two groups one has a higher elasticity, this group has a higher acceleration/lower deceleration of viral spread in comparison to the other one.

**Figure 1:**
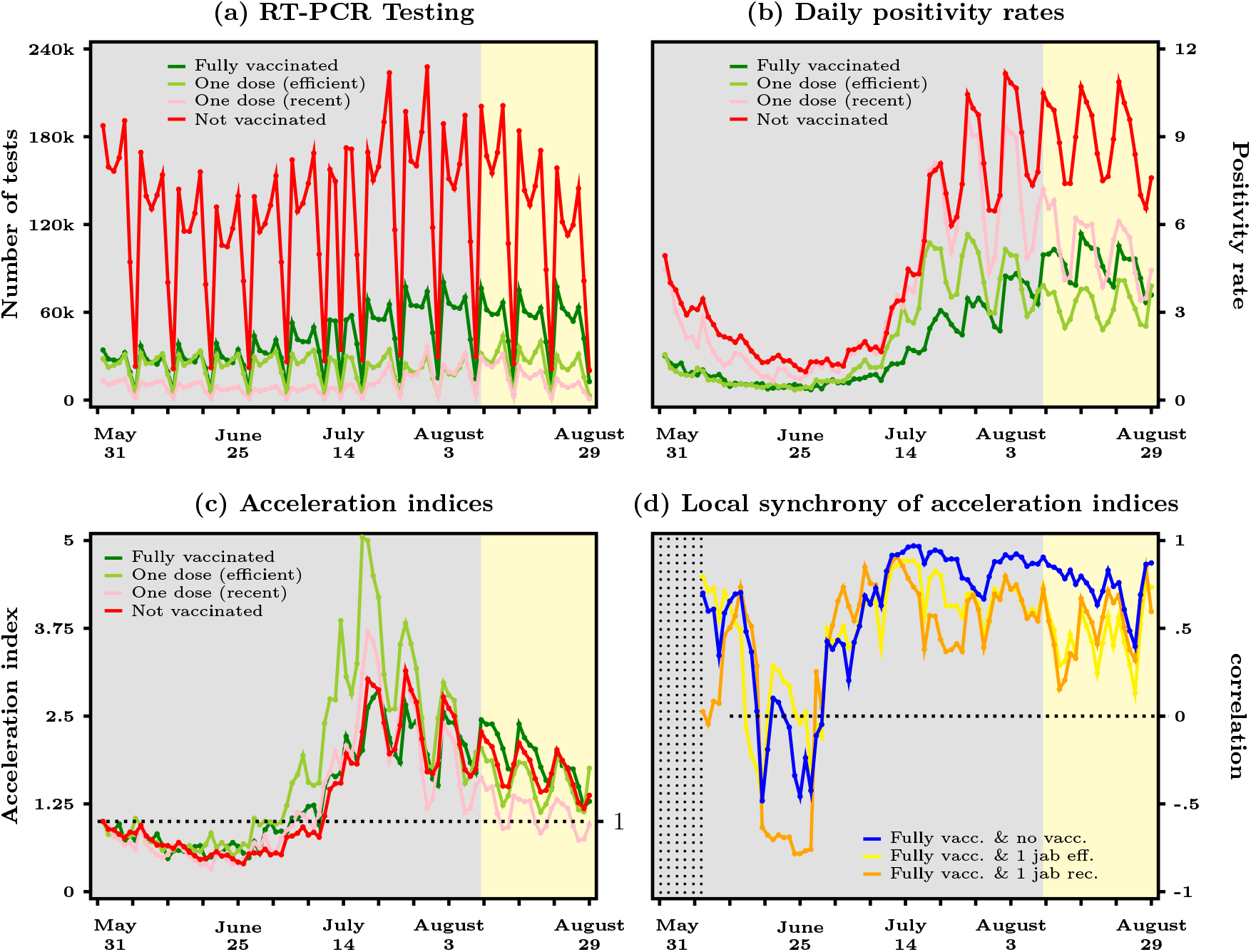
Epidemic dynamics in France across 4 groups with different vaccine status - May, 31 to August 29, 2021; panel (*a*) depicts the number of SARS-CoV-2 tests; panel (*b*) depicts positivity rates (defined as ratio of daily positive cases over daily tests); panel (*c*) depicts the acceleration indices (defined as the elasticity of cumulated cases to cumulated tests); panel (*d*) depicts Pearson correlation coefficients on a rolling window of 7 days between the acceleration index for the fully vaccinated and that of the other 3 groups; data obtained from DREES

This is exactly what the acceleration index does and in panel (*c*) we report its level for the 4 groups. Our acceleration index is lvel-free since, as we see in Equation (2), the daily positivity rate, i.e. the daily flows of positives and tests for a particular day, respectively *p* and *d*, are corrected by the average positivity rate, i.e. their stock values, respectively *P* and *D* for that particular day. We therefore see in panel (*c*) of Figure 1 that viral acceleration is essentially similar among fully vaccinated and unvaccinated individuals. This is because even though the unvaccinated have a larger daily positivity rate than the vaccinated, they also have a larger average rate: unvaccinated people are less protected against direct SARS-CoV-2 infection and therefore have a larger *P/D* over the epidemic past. But vaccinated people, who should be protected against direct SARS-CoV-2 infection have a lower daily positivity rate but with respect to a lower average positivity rate, which means that with respect to their epistemic past, viral spread is accelerating as quickly as among the non-vaccinated. This is insofar suprising as one may think that a succesful vaccination campaign would contribute to the deceleration of viral spread among the vaccinated. Yet this is not what we observe.

What is also striking in panel (*c*) is how close the acceleration indices stay over the entire period. More precisely, the resurgence of the acceleration regime, at the beginning of the summer break in early July, starts a little bit earlier for fully vaccinated individuals compared to unvaccinated people in France. The level of acceleration is about 1.2 for the two groups on August 29, 2021. This means with 1% of additional tests (the total number of test, or its stock, is increased by 1%), we find 1.2% additional cases (the total number of cases, or stock of positive cases, increases by 1.2%) in both groups. Acceleration for both groups reached its peak during the second half of July (a factor of about 3) and is steadily decreasing since the beginning of August. But the pandemic remains in an acceleration regime for both groups. The acceleration indices have been very much synchronized since the resurgence of the acceleration regime, starting early July. This is confirmed in panel (*d*), where we report the pairwise Pearson correlation coefficients on a rolling window of 7 days. In particular, the correlation between acceleration indices for fully vaccinated and unvaccinated individuals has reached its highest level mid-July and is still close to 1 at the end of the sample. In sum, our acceleration index shows no large difference across vaccinated and unvaccinated individuals since early/mid July.

## 4 Conclusion

The main result that we document in Figure 1 is that in France, COVID-19 acceleration has been of similar magnitude among both the vaccinated and the unvaccinated population since early July 2021, despite the daily positivity rate being larger among the latter group. Positivity rate is a level-dependent indicator that roughly informs us about viral spread or speed, but what is important to know is whether viral spread is accelerating or decelerating. Our acceleration index is a level-independent measure that indicates the percentage increase in cases following a percentage increase in tests and thus captures better whether the pandemic is under control or not. Our indicator can also serve as a basis for public health policy to curb COVID-19, that, if successful, would be one that sees ever fewer positive cases as testing efforts increase, i.e. our indicator would indicate deceleration. Currently, our indicator does not indicate deceleration of viral spread among the vaccinated, which may indicate that vaccination campaign is not fully achieving its aim.

The fact that acceleration persists even among the vaccinated population is broadly consistent with recent biological evidence that vaccinated and unvaccinated individuals have similar viral loads where the SARS-CoV-2 delta variant is most prevalent and that vaccine breakthroughs are seen more often in comparison with previous variants (see Brown et al, 2021; Musser et al, 2021; Riemersma et al, 2021; Public Health England Technical Briefing 20). Chia et al (2021) report that if vaccine breakthrough occurs, the viral load is the same for vaccinated and non-vaccinated individuals for the first few days, but that the viral load is decreasing more quickly for vaccinated individuals thereafter. A study from June 2021 in Vietnam has shown that the viral load with the delta variant in cases of vaccine breakthrough was over 250 times higher than in older strains (Chau et al. 2021). This means that while vaccination offers in general effective biological protection against severe cases and mortality (see e.g. Public Health England COVID-19 vaccination report 31; CDC Science Brief of September 15, 2021), it does not mean that vaccinated people cannot get infected with the virus and that they do not contribute to viral spread. Moreover, as it is known that even asymptomatic individuals may transmit the virus (e.g. Byambasuren, O. et al. 2020), asymptomatic vaccinated as well as non-vaccinated individuals may often go unperceived with no systematic testing effort. Testing for both, non-vaccinated *and* vaccinated individuals thus remains an important public health strategy to control the pandemic and to bring it into a deceleration regime. Indeed, research shows that test sensitivity is secondary to frequency for efficient COVID-19 screening (Larremore et al., 2021) and that there should not be a “one size fits all” testing strategy, primarily based on PCR-screening (Mina and Andersen 2021). The decision of the French authorities for example to concentrate mainly on vaccination to combat COVID-19, to make access Antigen-testing difficult and costly for individuals later this year, to focus on PCR-testing by prescription only and to impose a vaccine passport (“pass sanitaire”) that gives certain advantages to vaccinated people, notably to travel and to visit social gatherings without prior testing is questionable on the grounds that it does not guarantee to be effective to help curbing sustainably the COVID-19 acceleration that is at this date still observed in France.

## Data Availability

Data used in this note are available at https://data.drees.solidarites-sante.gouv.fr/explore/dataset/
covid-19-resultats-issus-des-appariements-entre-si-vic-si-dep-et-vac-si/information/?disjunctive.
vac_statut, accessed August 22, 2021.

https://data.drees.solidarites-sante.gouv.fr/explore/dataset/covid-19-resultats-issus-des-appariements-entre-si-vic-si-dep-et-vac-si/information/?disjunctive.vac_statut

## Footnotes

1 https://www.cdc.gov/coronavirus/2019-ncov/variants/delta-variant.html

2 See press releases by the DREES, in French, and notably the release on August 20, 2021 about differences in cases, tests and positivity rate between vaccinated and unvaccinated individuals, available at https://drees.solidarites-sante.gouv.fr/communique-de-presse/debut-aout-huit-fois-moins-de-tests-positifs-et-onze-fois-moins-dentrees-en, last accessed September 16, 2021.

3 Data used in this note are available at https://data.drees.solidarites-sante.gouv.fr/explore/dataset/covid-19-resultats-regionaux-issus-des-appariements-entre-si-vic-si-dep-et-vac-s/information/, accessed September 16, 2021.

## References

[1] Baunez C., Degoulet M., Luchini S., Pintus P., Teschl M. (2021): Tracking the Dynamics and Allocating Tests for COVID-19 in Real-Time: an Acceleration Index with an Application to French Age Groups and Départements. PLoS ONE, June 1st, available at https://doi.org/10.1371/journal.pone.0252443. 3, 4

[2] Blanquart, F., Abad, C., Ambroise, J., Bernard, M., Cosentino, G., Giannoli, JM., Débarre, F., (2021): Characterization of vaccine-breakthrough infections of SARS-CoV-2 Delta and Alpha variants and within-host viral load dynamics in the community in France. HAL-Working paper. Available at: https://hal.archives-ouvertes.fr/hal-03318483/

[3] Brown CM, Vostok J, Johnson H, et al. (2021): Outbreak of SARS-CoV-2 Infections, Including COVID-19 Vaccine Breakthrough Infections, Associated with Large Public Gatherings - Barnstable County, Massachusetts, July 2021. MMWR Morb Mortal Wkly Rep 2021;70:1059–1062, available at http://dx.doi.org/10.15585/mmwr.mm7031e2

[4] Byambasuren, O., Cardona, M., Bell, K., Clark, J., McLaws, ML., Glasziou, P. (2020): Estimating the extent of asymptomatic COVID-19 and its potential for community transmission: Systematic review and meta-analysis. JAMMI 5(4), pp. 223–234, available at https://jammi.utpjournals.press/doi/10.3138/jammi-2020-0030.

[5] Centers for Disease Control and Prevention (CDC) (2021): Science Brief: COVID-19 Vaccines and Vaccination, updated Sept, 15, 2021. Available at: https://www.cdc.gov/coronavirus/2019-ncov/science/science-briefs/fully-vaccinated-people.html.

[6] Chau, N., Ngoc, N., Nguyet, L. (2021):Transmission of SARS-CoV-2 Delta Variant Among Vaccinated Healthcare Workers, Vietnam. SSRN working paper, available at https://papers.ssrn.com/sol3/papers.cfm?abstract_id=3897733

[7] Chia, Po Ying and Xiang Ong, Sean Wei and Chiew Calvin J and Ang, Li Wei and Chavatte, Jean-Marc and Mak, Tze-Minn and Cui, Lin and Kalimuddin, Shirin and Chia, Wan Ni and Tan, Chee Wah and Ann Chai, Louis Yi and Tan, Seow Yen and Zheng, Shuwei and Pin Lin, Raymond Tzer and Wang, Linfa and Leo, Yee-Sin and Lee Vernon J and Lye, David Chien and Young, Barnaby Edward (2021): Virological and serological kinetics of SARS-CoV-2 Delta variant vaccine-breakthrough infections: a multi-center cohort study, MedRχv working paper, available at https://www.medrxiv.org/content/10.1101/2021.07.28.21261295v1

[8] Gazit, S., Shlezinger R., Perez, G., et al. (2021): Comparing SARS-CoV-2 natural immunity to vaccine-induced immunity: reinfections versus breakthrough infections. MedRχv working paper, available at https://www.medrxiv.org/content/10.1101/2021.08.24.21262415v1

[9] Larremore, D., Wilder, B., Lester, E., et al. (2021): Test sensitivity is secondary to frequency and turnaround time for COVID-19 screening. Sciences Advances 7(1), available at https://advances.sciencemag.org/content/7/1/eabd5393

[10] Layan, M., Gilboar, M., Gonen, T., et al. (2021): Impact of BNT162b2 vaccination and isolation on SARS-CoV-2 transmission in Israeli households: an observational study. MedRχv working paper, available at https://www.medrxiv.org/content/10.1101/2021.07.12.21260377v1

[11] Li, B., Deng, A., Li, K. et al. (2021): Viral infection and transmission in a large well-traced outbreak caused by the Delta SARS-CoV-2 variant. MedRχv working paper, available at https://www.medrxiv.org/content/10.1101/2021.07.07.21260122v2

[12] Musser, J., Christensen, P., Olsen, R., Long, S., Subedi, S., Davis, J., Hodjat, P., Walley, D., Kinskey, J., Gollihar, J. (2021): Delta variants of SARS-CoV-2 cause significantly increased vaccine breakthrough COVID-19 cases in Houston, Texas MedRχv working paper, available at https://www.medrxiv.org/content/early/2021/08/01/2021.07.19.21260808

[13] Olliaro, P., Torreele, E., Vaillant, M. (2021): COVID-19 vaccine efficacy and effectivenessâthe elephant (not) in the room. The Lancet 2(7), pp. E279–E280. Available at https://www.thelancet.com/journals/lanmic/article/PIIS2666-5247(21)00069-0/fulltext

[14] Planas, D., Veyer, D., Schwartz, O. et al. (2021): Reduced sensitivity of SARS-Cov-2 variant Delta to antibody neutralization. Nature 596, pp. 276–280. Available at https://www.nature.com/articles/s41586-021-03777-9

[15] Prunas, O., Warren, J., Crawford, F., et al. (2021): Vaccination with BNT162b2 reduces transmission of SARS-CoV-2 to household contacts in Israel. MedRχv working paper, available at https://www.medrxiv.org/content/10.1101/2021.07.13.21260393v1

[16] Public Health England Technical Briefing 20 (Aug 6, 2021): SARS-CoV-2 variants of concern and variants under investigationin England, available at https://assets.publishing.service.gov.uk/government/uploads/system/uploads/attachment_data/file/1009243/Technical_Briefing_20.pdf

[17] Public Health England COVID-19 Vaccination Report Week 31 (2021), available at https://assets.publishing.service.gov.uk/government/uploads/system/uploads/attachment_data/file/1008919/Vaccine_surveillance_report_-_week_31.pdf

[18] Puranik, A., Lenehan, P., Silvert, E., et al. (2021): Comparison of two highly-effective mRNA vaccines for COVID-19 during periods of Alpha and Delta variant prevalence. MedRχv working paper, available at https://www.medrxiv.org/content/10.1101/2021.08.06.21261707v3

[19] Riemersma, K., Grogan, B., Kita-Yarbro, A., Jeppson, G., O’Connor, D., Friedrich, T. Grande, K. (2021): Shedding of Infectious SARS-CoV-2 Despite Vaccination when the Delta Variant is Prevalent - Wisconsin, July 2021. MedRχv working paper, available at https://www.medrxiv.org/content/10.1101/2021.07.31.21261387v3.

[20] Salo, J., Hägg, M., Kortelainen, M. et al. (2021): The indirect effect of mRNA-based Covid-19 vaccination on unvaccinated household members. MedRχv working paper, available at https://www.medrxiv.org/content/10.1101/2021.05.27.21257896v2

[21] Uriu, K., Kimura, I., Shirakawa, K., et al. (2021): Ineffective neutralization of the SARS-CoV-2 Mu variant by convalescent and vaccine sera. BioRχv working paper, available at https://www.biorxiv.org/content/10.1101/2021.09.06.459005v1

